# CYP2C19 and effect of clopidogrel for secondary prevention of major ischemic events: systematic review and meta-analysis

**DOI:** 10.1101/2023.08.17.23292850

**Authors:** Femke C.C. Kremers, Jochem van den Biggelaar, Hester F. Lingsma, Ron H.N. van Schaik, Bob Roozenbeek, Diederik W.J. Dippel

**Author notes:** **Corresponding Author: Femke Kremers**.

## Abstract

**Background:** Carriers of the CYP2C19 Loss of Function (LoF) allele may experience decreased efficacy of clopidogrel in secondary prevention after cardiovascular events. Randomized clinical trials of clopidogrel in patients with known CYP2C19 carrier status provided inconsistent results. Our aim was to pool the evidence on the effect of clopidogrel on outcome, according to CYP2C19 LoF status, in a meta-analysis.

**Methods:** A systematic review and meta-analysis of randomized controlled trials (RCTs) on the effect of clopidogrel according to CYP2C19 LoF status in patients with cardiovascular disease and recent TIA or stroke was performed. The primary outcomes were 1) ischemic stroke and 2) major adverse cardiac events (MACE). We used random effects analysis to estimate the effect of clopidogrel as a pooled odds ratio (OR) in carriers and non-carriers of the LoF variant and tested for interaction between clopidogrel and CYP2C19.

**Results:** We included six RCTs with a total of 15,141 participants comparing combined clopidogrel and aspirin therapy to aspirin monotherapy. The effect of clopidogrel on MACE was OR=0.70 in CYP2C19 non-variant carriers compared to OR=0.84 in CYP2C19 variant carriers (pinteraction=0.13). In patients with a recent TIA or minor ischemic stroke, the OR for the effect of clopidogrel on MACE was OR=0.52 in CYP2C19 non-variant carriers compared to OR=0.84 in CYP2C19 variant carriers (pinteraction=0.02) while in patients with cardiovascular disease the difference in effect was not evident (non-variant carriers OR=0.76, variant carriers OR= 0.84, pinteraction=0.50).

**Conclusion:** Our meta-analysis could not establish that overall, clopidogrel is less effective in patients with a recent MI, minor stroke or TIA and a CYP2C19 LoF genotype. However, the size and direction of the difference in effect warrants further research.

**Registration - URL**: https://www.crd.york.ac.uk/prospero/; Unique identifier: CRD42021242993.

## Introduction

Patients with a recent transient ischemic attack (TIA), minor ischemic stroke (IS) or cardiac disease, including acute coronary syndrome (ACS) and atrial fibrillation (AF), are at increased risk of recurrent ischemic events.^1, 2^

Recurrent ischemic events are not uncommon in these high-risk patients, even when they are treated with antiplatelet agents, such as clopidogrel. This may partly be ascribed to the incomplete efficacy of these therapies. Another possible cause for the high numbers of recurrent ischemic events is that some patients are unable to convert the prodrug of clopidogrel to its active metabolite. This conversion is regulated by the hepatic cytochrome p450 enzymes, of which hepatic cytochrome p450 2C19 (CYP2C19) is an important contributor.^3^

Multiple variant alleles of the *CYP2C19* gene are associated with different levels of activity of the CYP2C19 enzyme. The *17 allele variant (-806T>C; rs12248560) is associated with an increased CYP2C19 activity (Gain-of-function), whereas the *2 (681G>A; rs4244285) and *3 (636G>A; rs4986893) variants are loss-of-function (LoF) alleles and produce a CYP2C19 enzyme with decreased activity. Individuals carrying *2 or *3 alleles are called ‘LoF-carriers’ and have a reduced clopidogrel metabolism.^4^ These LoF genetic variants occur in approximately 30% of the Western population and in up to 60% in the Asian population.^3, 5, 6^

The debate on the utility and effectiveness of testing for this purpose is ongoing and several testing strategies have been proposed and described in observational studies. Direct evidence of the interaction of CYP2C19 variants with clopidogrel on recurrent vascular events is not abundantly available.^7^ In the randomized controlled trials that investigated whether clopidogrel + ASA is superior to ASA monotherapy for the prevention of new ischemic events, genetic sub-studies investigated the effect of *CYP2C19* LoF variants on the effect of clopidogrel treatment.^5, 8, 9^ The results of genetic sub-studies differ, and several studies were not sufficiently powered to detect a difference in effect between subgroups of patients with *CYP2C19* LoF variants.^7^

To create clarity on this subject, we performed a systematic review and meta-analysis to estimate the effect of the *CYP2C19* LoF variants on the effect of clopidogrel treatment in patients with either cardiac ischemic disease or a recent TIA or minor ischemic stroke.

## Methods

### Search strategy

We performed a comprehensive literature search in the online databases Embase, Medline ALL, Web of Science Core Collection, Cochrane Central Register of Controlled Trials (CENTRAL) and Google Scholar on March 13, 2023 to include randomized controlled trials (RCT) that investigated patients with cerebrovascular disease or cardiovascular disease treated with clopidogrel versus without clopidogrel and were followed for the occurrence of IS, major bleeding or a composite outcome for major cardiovascular events (MACE) of IS, myocardial infarction (MI), non-central nervous system systemic embolism or vascular death (Supplementary Material I). In the present meta-analysis, we also included trials in patients with cardiovascular disease, such as acute coronary syndrome (ACS), peripheral artery disease (PAD), atrial fibrillation (AF) and patients with multiple cardiovascular risk factors, who are at increased risk of recurrent ischemic events. CYP2C19 status should have been investigated in the trial population by genotyping extracted DNA of the participants.

This systematic review was registered with PROSPERO, the International Prospective Registry of Systematic Reviews (CRD42021242993).^10^ We performed this systematic review in accordance with the Preferred Reporting Items for Systematic Review and Meta-Analysis (PRISMA) guidelines,^11^ and we filled out the checklist (Supplementary Table I).

### Outcomes

The investigated outcomes were 1) occurrence of fatal or non-fatal ischemic stroke or 2) a major cardiovascular event (MACE; IS, MI, non-central nervous system embolism or vascular death). The safety outcome was a major bleeding event, defined as symptomatic intracranial hemorrhage, intraocular hemorrhage causing vision loss, or other bleeding resulting in substantial hemodynamic compromise requiring treatment.^12^

### Study selection

Two assessors (JB and FK) independently screened the retrieved articles by title and abstract to identify studies that met the inclusion criteria. Both assessors had to agree with inclusion after screening the eligible studies based on title and abstract. When discrepancies occurred with full text screening of the articles, the discrepancy was discussed with both assessors until consensus was reached. If multiple publications were based on the same trial, the publication with the most complete dataset was included.

### Quality assessment and data extraction

Relevant data were extracted from each trial included in the meta-analysis. The following data were extracted: authors, year of publication sub study, enrollment, participants, ancestry, number participants in sub study, reported LoF-carriers, *CYP2C19* variants, median follow-up period, outcomes, safety outcome, treatment, blinding.

The two assessors (JB and FK) separately evaluated the risk of bias of each included trial by using the Cochrane Collaboration Risk of Bias assessment tool.^13^ The following domains were assessed: random sequence generation, allocation concealment, blinding of participants and personnel, blinding outcome assessment, incomplete outcome data, selective outcome reporting, and other sources of bias. Items were scored as ‘low risk of bias’, ‘some concerns’, or ‘high’ risk of bias (Supplementary Material II).

### Statistical analysis

Based on the reported numbers of outcomes (IS, major bleeding and MACE) in the intervention and control groups we estimated the effect of clopidogrel in each study. The pooled effects in the CYP2C19 groups were estimated with a random effects model. The p-value for interaction was estimated by including an interaction factor between treatment and *CYP2C19* variant subgroup in the random effects model. The interaction was considered statistically significant at p<0.05.

All results are reported as odds ratios (OR) with 95% confidence intervals (CI) and will be presented in forest plots. Heterogeneity was assessed with the Cochran’s *Q* test and the *I*^2^ statistic to evaluate the variance between the studies per outcome for carriers and non-carriers separately. The Cochran’ Q test was considered statistically significant at p<0.1. Analyses were stratified for the cardiovascular disease population and the TIA or minor stroke population. Sensitivity analyses were performed by considering “low risk of bias” studies only.

Statistical analyses were performed with R Studio statistical software version 1.3.1093 for the primary outcome and safety outcome. The following packages were used in the analysis: ‘glm2’, ‘readxl’, ‘lme4’, ‘meta’, ‘metafor’ and ‘forestplot’.

## Results

### Study selection

The search provided 1233 articles for inclusion. After screening on title and abstract, 131 articles were left for full text assessment. Conference abstracts were excluded. Studies with genotype-guided treatment allocation and trials that did not investigate the required treatment contrast were excluded after full text assessment. Trials comparing clopidogrel with a different P2Y12-receptor inhibitor such as ticagrelor or prasugrel were excluded. ASA exerts its anti-platelet effect through cyclooxygenase (COX) inhibition instead of the P2Y12-receptor and was therefore accepted as co-medication in the trials. Publications based on the same clinical trial led to exclusion of the least complete publication. In the end, 5 eligible articles describing 6 RCTs were left for inclusion in the meta-analysis (Figure 1, Table I, Supplementary Table III). ^5, 8, 14-16^ These included a total of 15,137 patients of which 5,260 were genotyped with at least one *2 or *3 *CYP2C19* LoF variant allele (Table 1, Supplementary Table II, Supplementary Table III). Publication bias was low in included studies (Figure 2).

**Figure 1:**
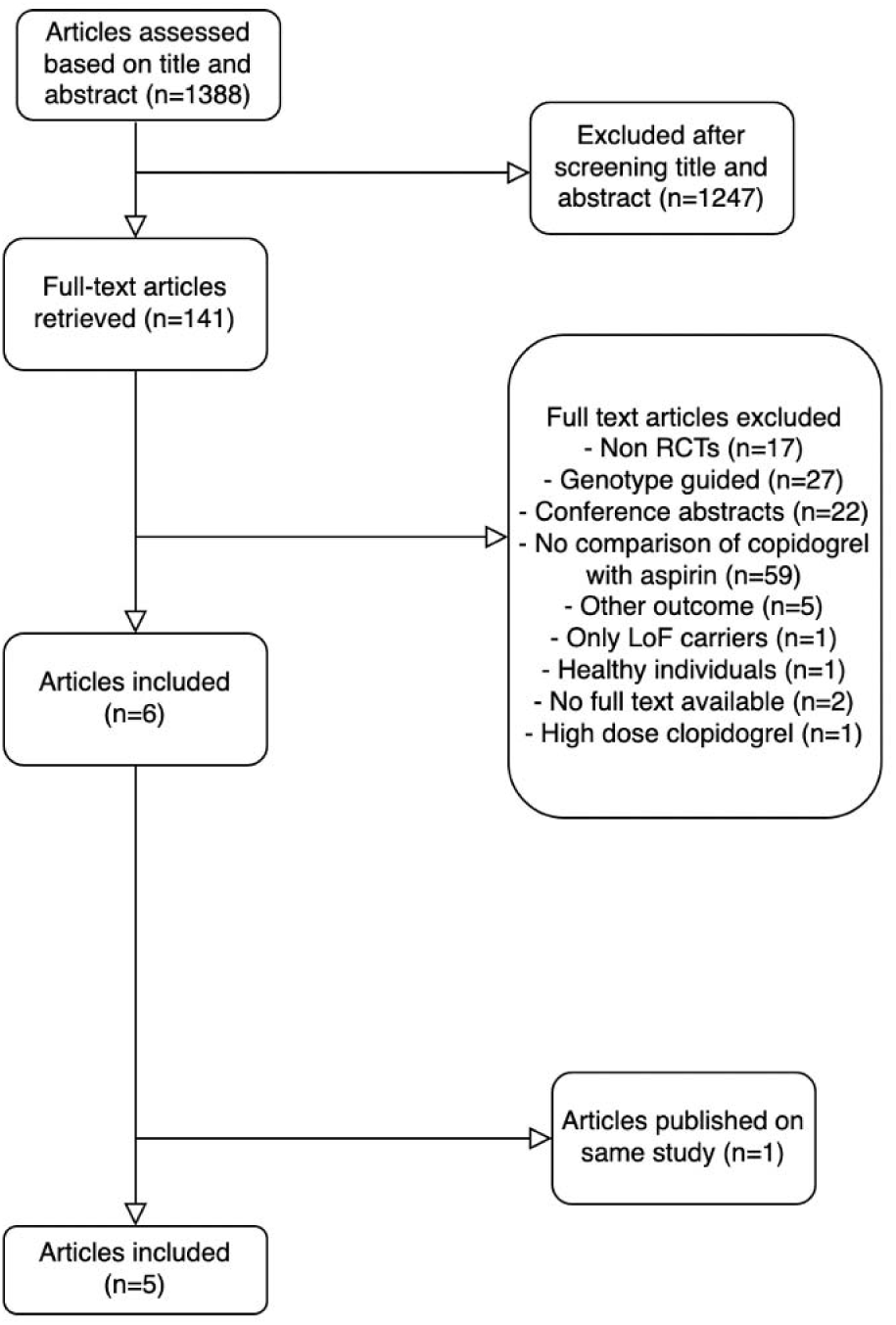
Flow diagram of selection procedure.

**Figure 2:**
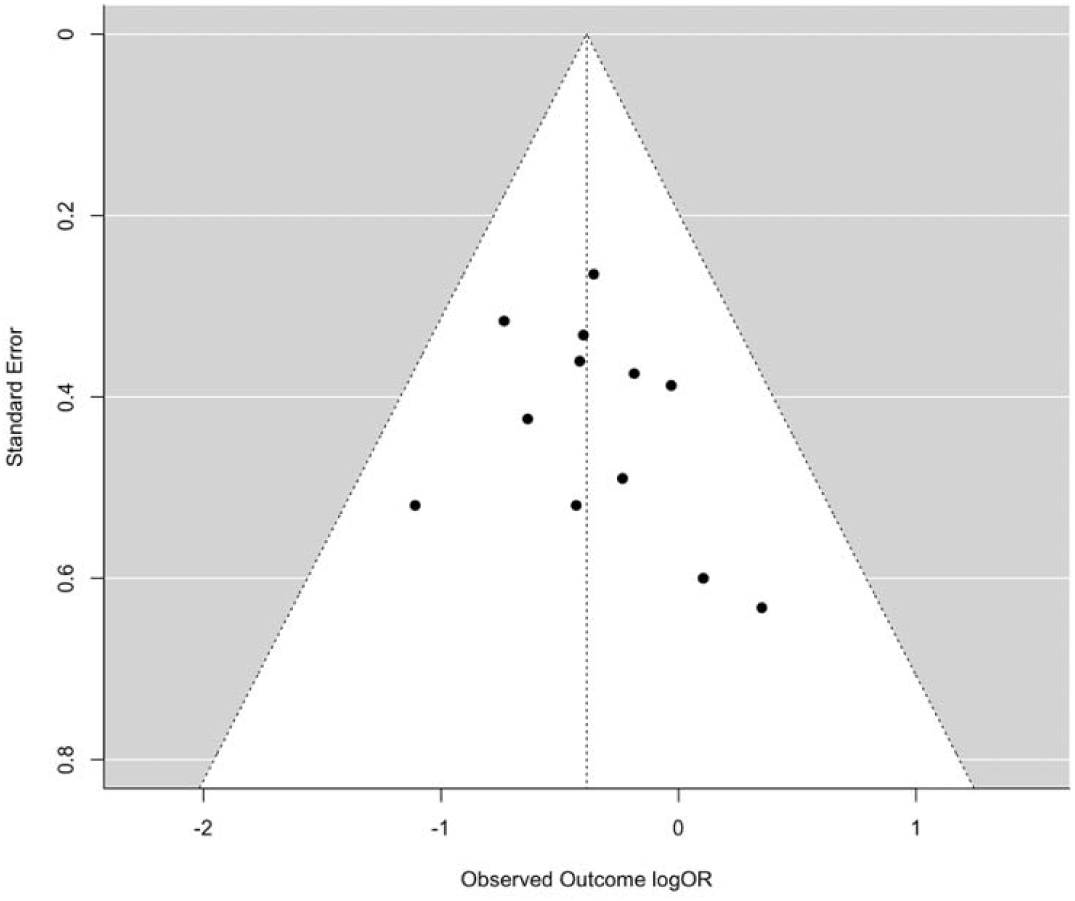
Funnel plot for publication bias in included studies.

**Table 1:**
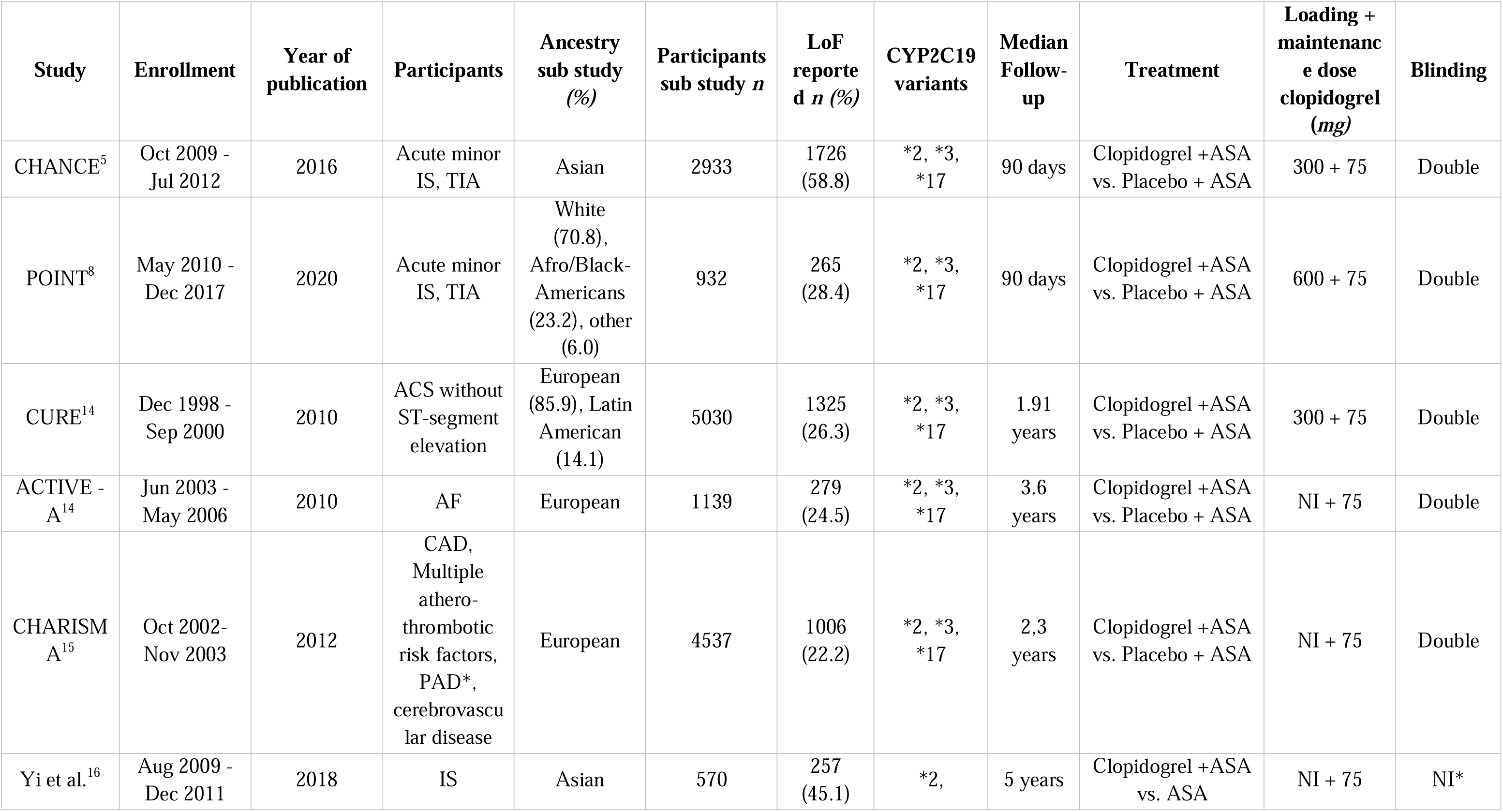
Study characteristics of the included trials.

### Study characteristics

In three studies, the index event was a TIA or minor stroke^5, 8, 16^, while three other studies included patients with cardiovascular disease. ^14, 15^ Two studies were executed in Asian countries,^5, 16^ the other four were mainly based in Europe or North America.^8, 14, 15^ All studies assessed major vascular events (MACE),^5, 8, 14-16^ and five studies reported major bleeding events. ^5, 8, 14, 15^

The intervention group of all included trials received clopidogrel (75 mg daily) + ASA. In three studies, participants in the intervention group received a loading dose of clopidogrel varying from 300 to 600 mg. ^5, 8, 14^ The control group received ASA + placebo in five studies, in one study the control group only ASA without placebo (Supplementary Table III). ^16^ The median follow-up period was 90 days in two trials of early antiplatelet treatment. ^5, 8^ The median follow-up period varied from 1,9 years to 5 years in the other studies. ^14-16^

### Quality assessment

Risk of bias was low in all included trials for the following domains: random sequence generation, allocation concealment, blinding outcome assessment, incomplete outcome data, and other sources of bias. The domains reporting outcome data and blinding of personnel and participants were assessed as ‘some concern’ in one study.^16^ For that reason, the overall risk of bias judgement for this trial was ‘some concerns’, all other included trials were scored as ‘low risk’ (Supplementary Table IV).

### Effect of clopidogrel

Two trials assessed IS separately from the composite outcome of MACE, both trials consisted of TIA or minor stroke patients ^5, 8^. The observed effect of clopidogrel on IS was larger in non-carriers (OR:0.51, 95%CI: 0.36-0.73) than in carriers (OR: 0.79, 95%CI: 0.58 - 0.1.08) without a significant interaction effect (p=0.07) (Figure 3). No significant heterogeneity was measured for carriers (p=0.17) and noncarriers (p=0.56).

**Figure 3:**
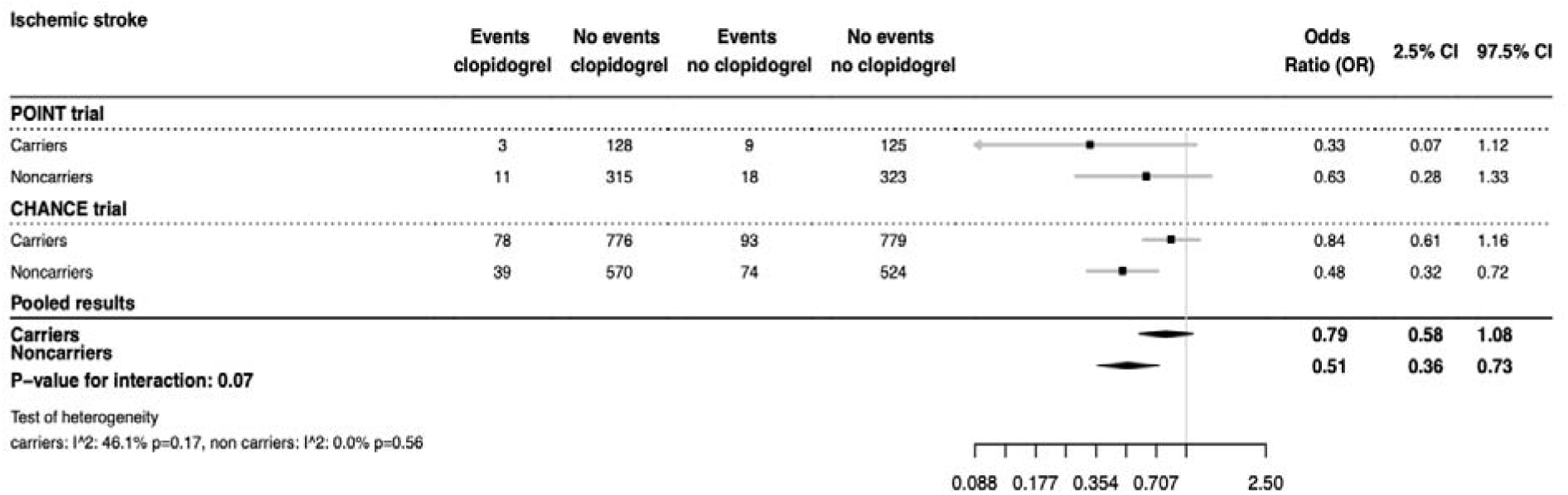
Effect of *CYP2C19* variant and occurrence of ischemic stroke.

The effect of clopidogrel for the total population on MACE was larger in non-carriers (OR: 0.70, 95%CI: 0.62-0.80) than in carriers (OR: 0.84, 95%CI: 0.70-1.02), but without significant interaction (p=0.13) (Figure 4). For MACE, carriers showed small to moderate heterogeneity (*I*^2^=39.18%, p=0.14), while noncarriers showed moderate heterogeneity (*I*^2^=43.42%, p=0.12) (Figure 4). In TIA or minor stroke patients, the effect of clopidogrel on MACE was larger in non-carriers (OR:0.52, 95%CI: 0.38-0.71) than in carriers (OR: 0.84, 95%CI: 0.64-1.11), with a significant interaction (p=0.02) (Figure 4). The effect of clopidogrel on MACE in the cardiovascular subgroup was larger in non-carriers (OR: 0.76, 95%CI:0.65-0.88) than in carriers (OR: 0.84, 95%CI: 0.64-1.09) but the difference was smaller and not significant (p=0.50) (Figure 4).

**Figure 4:**
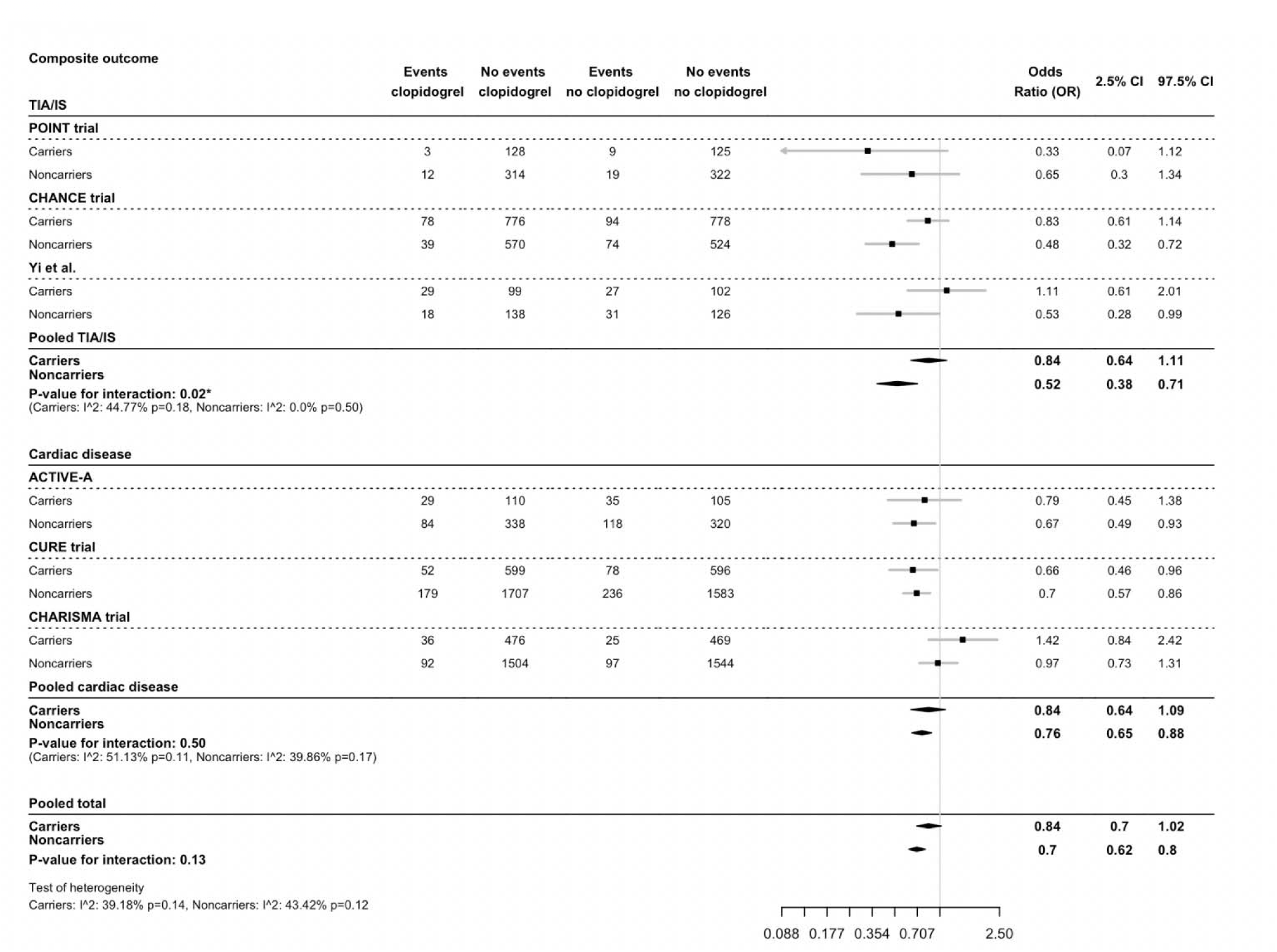
Effect of *CYP2C19* variant and treatment on major vascular events.

The effect of clopidogrel on major bleeding risk was smaller in non-carriers (OR:1.27, 95%CI: 1.00-1.61) than in carriers (OR: 2.48, 95%CI: 1.57-3.83), with a significant interaction (p=0.01) (Figure 5). No significant heterogeneity was measured (*I*^2^=16.62%, p=0.31).

**Figure 5:**
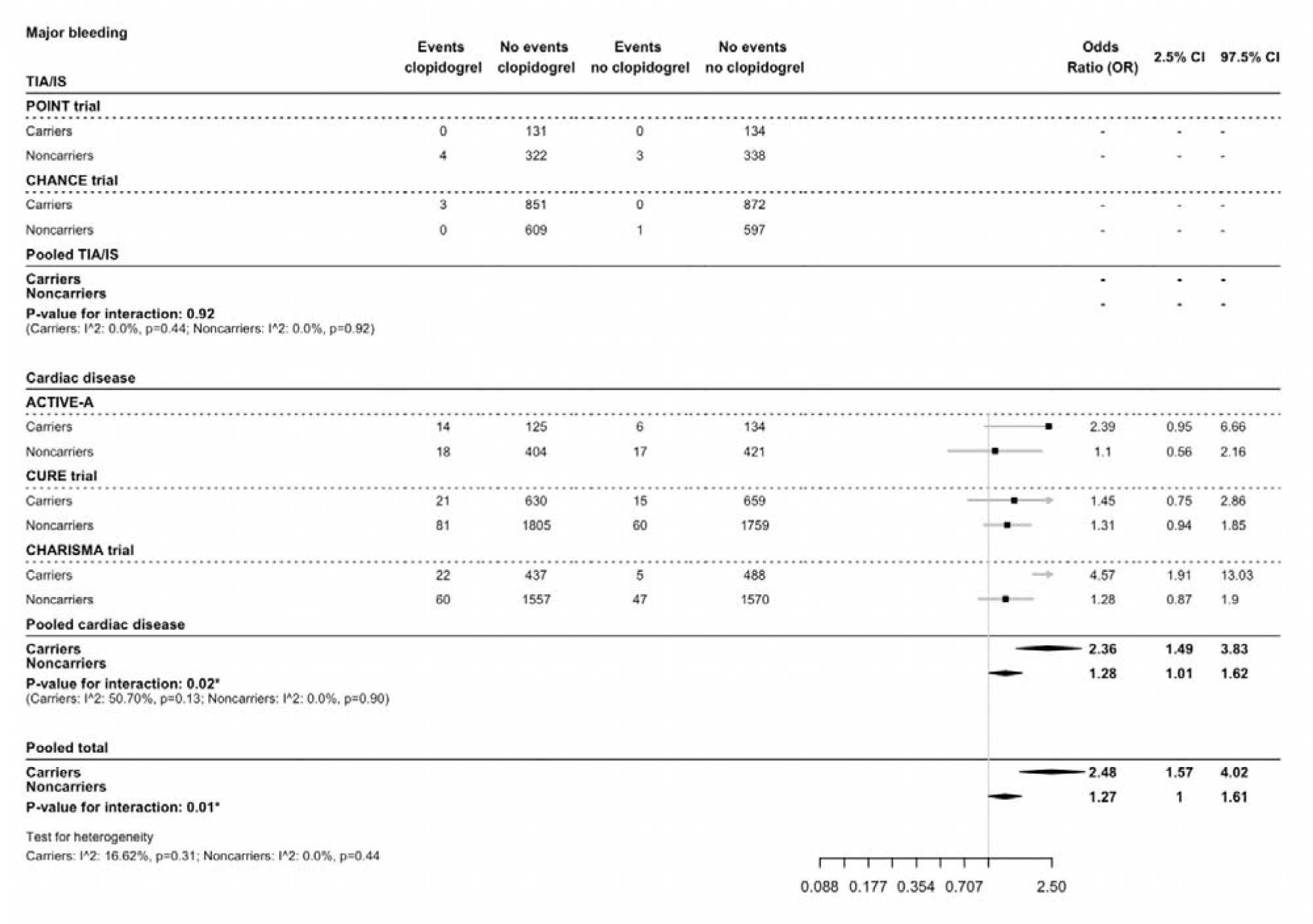
Effect of *CYP2C19* variant and treatment on major bleeding.

In the cardiovascular disease population, the effect of clopidogrel treatment was smaller in non-carriers (OR: 1.28, 95%CI:1.01-1.62) than in carriers (OR:2.36, 95%CI:1.49-3.83) with an interaction of p=0.02 (Figure 5). Because of the low number of bleeding events in the included trials, the effect of clopidogrel on major bleeding in patients with TIA or minor stroke could not be estimated.

In the sensitivity analysis with only low risk of bias trials included, the results were consistent, although the p-value in the TIA or minor stroke population for MACE was not significant (p = 0.08).

## Discussion

In this systematic review and meta-analysis, we evaluated how *CYP2C19* LoF allelic variants influence the effect of clopidogrel on the occurrence of major vascular events or major bleeding in patients with a recent TIA, minor stroke or cardiovascular disease. After combining data from 6 studies in patients with TIA, minor IS or cardiovascular disease, we found a reduction of effect of clopidogrel in CY2C19 variantcarriers with an OR of 0.84 (95%CI 0.7-1.02) compared to 0.70 (95%CI 0.62-0.80) in non-carriers (p_interaction_=0.13). In the subgroup of patients with ischemic stroke, the risk of MACE was significantly higher compared to non-carriers (0.84 (95%CI 0.64-1.11 vs. 0.52, 95%CI 0.38-0.71, p_interaction_=0.02).

The protective effect of clopidogrel on the occurrence of MACE may be diminished in patients with TIA or minor stroke due to the *CYP2C19* LoF variant but our results are not decisive. The interaction of the *CYP2C19* LoF variant with clopidogrel on the occurrence of ischemic stroke seemed less in carriers, but could not be proven since the interaction was not statistically significant. The interaction of the *CYP2C19* LoF variant with clopidogrel effect on MACE was not also statistically significant overall, and the significant interaction of effect of clopidogrel on MACE in studies of patients with TIA or minor stroke may represent a type 1 error in an overall neutral analysis.

The direction of the interaction was however consistent in the two major subgroups. The effect of clopidogrel by carrier status was significant in patients with a recent TIA or minor stroke, but may have been a type I error caused by multiplicity. In addition, and rather surprisingly, the risk of major bleeding was significantly higher in carriers of the *CYP2C19* allele compared to non-carriers when on clopidogrel treatment.

The association between clopidogrel and *CYP2C19* LoF variants has been investigated in other settings.^17-19^ Some studies have performed a meta-analysis to assess the influence of *CYP2C19* LoF variants in patients with a history of cardiovascular disease only. Only one meta-analysis assessed influence of *CYP2C19* LoF variants on TIA and minor stroke patients.^9^ In these studies, the beneficial effect of clopidogrel treatment on ischemic events in patients with a LoF variant was smaller than in patients without a LoF variant. These meta-analyses included randomized controlled trials, case-control studies and cohort studies, which makes them difficult to interpret. In our study, we only included randomized controlled trials, in order to minimize confounding and selection bias. In addition, most of the published meta-analyses did not assess interaction to assess the unbiased effect of a *CYP2C19* LoF variant on clopidogrel efficacy with random effects modelling.

Our study population consisted of participants with different manifestations of vascular disease and therefore different pathophysiological mechanisms leading to ischemic events. Clopidogrel has platelet aggregation lowering properties through inhibition of the P2Y12-receptor on platelets, to prevent thrombus formation in the blood vessels. Compared to cardiac ischemia, a minor ischemic stroke or TIA is more often caused by a blockage of the blood vessel with a thrombo-embolus. ^20^ Cardiac ischemia, on the other hand, is more often caused by reduced coronary artery perfusion due to local atherosclerosis. ^21, 22^ The difference in pathophysiology and the point of application of the active substances of clopidogrel may explain why no significant interaction effect was found in the cardiac disease population, although a significant relationship between *CYP2C19* LoF variants and clopidogrel effectiveness has been observed in other studies for patients with cardiac disease. ^23^

In addition to the available literature on the relationship of *CYP2C19* LoF variants and clopidogrel efficacy, multiple studies have been performed to investigate the implementation of genotype-guided therapy. ^23-28^ These studies have mainly been performed for patients with a history of cardiovascular disease ^26, 29^ and they show promising results regarding implementation of genotype-guided treatment but were not conclusive. ^23, 30-32^ In the CHANCE-2 trial, minor stroke patients with a *CYP2C19* LoF mutation showed better response on treatment with ticagrelor and aspirin compared to treatment with clopidogrel and aspirin. ^28^

More studies on genotype-guided treatment for patients with cardiovascular disease as well as patients with a recent TIA or minor stroke and the implementation in real clinical practice are needed.

### Limitations

Our study has several limitations. All studies were performed as a post-hoc analysis. Patients were genotyped for inclusion in a genetic sub-study of the original trial. In most studies, the genotype had been determined in only a small proportion of patients from the original trial. Only one fifth of the participants in the POINT and ACTIVE-A trial were genotyped for *CYP2C19* LoF variants.^8, 14^ However, as the trials were all randomized, selection and confounding bias is unlikely. Third, in the subgroup of trials assessing clopidogrel effect in patients with a TIA or minor stroke, 60% of all events were from the CHANCE trial. The number of events was considerably smaller in the two remaining trials (Yi et al., POINT). Therefore, the CHANCE trial has played a major role in the effect found for the TIA/minor stroke population. The absence of a differential effect of CYP2C19 in the POINT trial may have been caused by the higher clopidogrel loading dose of 600 mg that was employed. ^33, 34^ In the POINT trial, most events occurred within the first 48h after randomization, which is consistent with earlier studies.^1, 2^ This higher loading dose could have prevented more events in the first days compared to the CHANCE trial, which could be one of the causes of the difference between the results of these trials. A higher dose of clopidogrel maintenance treatment may influence the effectiveness on patients with 1 loss-of function allele.

Studies consisted of heterogenous patient populations, since the outcome of a cardiac event differed per study. Although we performed separated analyses according to the population, the comparability of the populations may limit the interpretation of the findings. In the CURE study, patients with acute coronary syndrome were included, while in the ACTIVE-A patients with atrial fibrillation were analyzed. In the CHARISMA trial, a subgroup of TIA/IS patients were also included in the patient population.

Patients with one *CYP2C19* LoF allele and one functioning allele are considered intermediate metabolizers, whereas poor metabolizers have very low conversion of clopidogrel to its active metabolite by CYP2C19. When patients have suboptimal conversion of clopidogrel and therefore less effective platelet inhibition, these patients may benefit from a higher loading and maintenance dose of clopidogrel. ^35, 36^ However, no consensus exists on the effectiveness of a higher maintenance dose of clopidogrel for patients with a *CYP2C19* LoF variant allele.^37, 38^

In the present study, only carriers vs. noncarriers of the *CYP2C19* allele were analyzed. We did not separately investigate poor and intermediate metabolizers of CYP2C19, since the number of events for each outcome within the poor metabolizer group was very small in all included studies. The investigated patient population mainly consisted of intermediate metabolizers (Table 1).

In the POINT trial, baseline National Institutes of Health Stroke Scale (NIHSS) for minor ischemic stroke or TIA patients was 2.^8^ In the CHANCE trial, the baseline NIHSS score was less than 3 for inclusion, and in the article of Yi et al, this was approximately 11 points.^5, 16^ Since in the 2 largest trials (CHANCE and POINT) the baseline NIHSS was low, caution is needed to extrapolate these results in real clinical practice, where patients often experience more disabilities at presentation.

We assessed safety outcomes as events of major bleeding for both patients with cardiovascular disease and patients with a history of TIA or minor stroke. Unfortunately, we could not assess the effect of CYP2C19 on bleeding events for patients with TIA or minor stroke due to a small number of events, which made it statistically inaccurate to interpret these results. For patients with cardiovascular disease and for the overall pooled results, the adverse effect of bleeding due to clopidogrel treatment was higher in patients with a *CYP2C19* LoF variant allele compared to patients without a LoF variant. This finding is not in agreement with earlier studies which showed lower bleeding rates in patients with a *CYP2C19* LoF variant. ^9, 39, 40^ More research is needed to assess the effect of a LoF variants on bleeding events in patients with TIA, minor stroke or cardiovascular disease.

Recent AHA or ESC guidelines do not address CYP2C19 genotyping, nor does the ESO guideline on antithrombotic treatment for stroke prevention. ^41-45^ Future studies regarding cost-effectiveness and effect in real life settings of *CYP2C19* genotype guided testing may provide insight in its prevention of recurrent ischemic events.

## Conclusion

Our meta-analysis could not establish that overall, clopidogrel is less effective in patients with a recent MI, minor stroke or TIA and a *CYP2C19* LoF genotype. However, the size and direction of the difference in effect warrants further research into the role of LoF genotypes and the cost-effectiveness of testing.

## Data Availability

All data is derived from published literature.

## Nonstandard Abbreviations and Acronyms

MACE: Major Cardiovascular Event
TIA: Transient Ischemic Attack
LoF: Loss-of-function
IS: Ischemic Stroke
ASA: Aspirin

## Supplemental Materials

Supplementary Material I, II

Supplementary Tables I – IV

