## Supplemental files for "CYP2C19 and effect of clopidogrel for secondary prevention of major ischemic events: systematic review and meta-analysis"

**Supplementary Material I.** Literature search

| Database searched | via | Years of coverage | Records | Records after duplicates removed |
|---|---|---|---|---|
| Embase | Embase.com | 1971 - Present | 868 | 849 |
| Medline ALL | Ovid | 1946 - Present | 435 | 46 |
| Web of Science Core Collection | Web of Knowledge | 1975 - Present | 681 | 286 |
| Cochrane Central Register of Controlled Trials | Wiley | 1992 - Present | 334 | 77 |
| Other sources: Google Scholar | | | 200 | 130 |
| **Total** | | | **2518** | **1388** |

**Embase.com**

('clopidogrel'/de OR 'antithrombocytic agent'/de OR 'purinergic P2Y receptor antagonist'/de OR 'dual antiplatelet therapy'/de OR (clopidogrel* OR Plavix* OR P2Y12 OR ((antiplatelet* OR anti-platelet* OR antithrombocyt* OR anti-thrombocyt*) NEXT/1 (therap* OR treatment*))):ab,ti,kw) AND ('brain ischemia'/exp OR 'coronary artery disease'/exp OR 'acute coronary syndrome'/exp OR 'ischemic heart disease'/exp OR 'major adverse cardiac event'/de OR 'cerebrovascular accident'/exp OR 'bleeding'/exp OR 'stroke patient'/exp OR 'cardiovascular disease'/de OR 'cerebrovascular disease'/de OR (stroke* OR TIA OR acute-coronar*-syndrome* OR ((coronary) NEAR/3 (arter* OR heart*) NEAR/3 (disease*)) OR ((cardiovascular* OR cardiac*) NEAR/3 (event*)) OR bleeding* OR ((cerebrovascular) NEAR/3 (accident* OR disease*)) OR ischemi* OR (myocard* NEAR/3 infarct*)):ab,ti,kw) AND ('cytochrome P450 2C19'/de OR (CYP2C19* OR CYP-2C19* OR cytochrome-P450-2C19*):ab,ti,kw) AND ('Controlled clinical trial'/exp OR 'Crossover procedure'/de OR 'Double-blind procedure'/de OR 'Single-blind procedure'/de OR (random* OR factorial* OR crossover* OR (cross NEXT/1 over*) OR placebo* OR ((doubl* OR singl*) NEXT/1 blind*) OR assign* OR allocat* OR volunteer* OR trial OR groups):ab,ti,kw) NOT ((animal/exp OR animal*:de OR nonhuman/de) NOT ('human'/exp))

**Medline (Ovid)**

(Clopidogrel / OR Purinergic P2Y Receptor Antagonists/ OR Dual Anti-Platelet Therapy/ OR (clopidogrel* OR Plavix* OR P2Y12 OR ((antiplatelet* OR anti-platelet* OR antithrombocyt* OR anti-thrombocyt*) ADJ1 (therap* OR treatment*))).ab,ti,kf.) AND (Cerebrovascular Disorders/ OR exp Brain Ischemia/ OR Coronary Artery Disease/ OR exp Myocardial Ischemia/ OR exp Stroke/ OR exp Hemorrhage/ OR Cardiovascular Diseases/ OR (stroke* OR TIA OR acute-coronar*-syndrome* OR ((coronary) ADJ3 (arter* OR heart*) ADJ3 (disease*)) OR ((cardiovascular* OR cardiac*) ADJ3 (event*)) OR bleeding* OR ((cerebrovascular) ADJ3 (accident* OR disease*)) OR ischemi* OR (myocard* ADJ3 infarct*)).ab,ti,kf.) AND (Cytochrome P-450 CYP2C19/ OR (CYP2C19* OR CYP-2C19* OR cytochrome-P450-2C19*).ab,ti,kf.) AND (exp Controlled clinical trial/ OR "Double-Blind Method"/ OR "Single-Blind Method"/ OR "Random Allocation"/ OR (random* OR factorial* OR crossover* OR cross over* OR placebo* OR ((doubl* OR singl*) ADJ blind*) OR assign* OR allocat* OR volunteer* OR trial OR groups).ab,ti,kf.) NOT (exp animals/ NOT humans/)

**Web of Science**

TS=(((clopidogrel* OR Plavix* OR P2Y12 OR ((antiplatelet* OR anti-platelet* OR antithrombocyt* OR anti-thrombocyt*) NEAR/1 (therap* OR treatment*)))) AND ((stroke*

OR TIA OR acute-coronar*-syndrome* OR ((coronary) NEAR/2 (arter* OR heart*) NEAR/2 (disease*)) OR ((cardiovascular* OR cardiac*) NEAR/2 (event*)) OR bleeding* OR ((cerebrovascular) NEAR/2 (accident* OR disease*)) OR ischemi* OR (myocard* NEAR/2 infarct*))) AND ( (CYP2C19* OR CYP-2C19* OR cytochrome-P450-2C19*)) AND ((random* OR factorial* OR crossover* OR (cross NEAR/1 over*) OR placebo* OR ((doubl* OR singl*) NEAR/1 blind*) OR assign* OR allocat* OR volunteer* OR trial OR groups)))

**Cochrane Central**

((clopidogrel* OR Plavix* OR P2Y12 OR ((antiplatelet* OR anti NEXT platelet* OR antithrombocyt* OR anti NEXT thrombocyt*) NEXT/1 (therap* OR treatment*))):ab,ti,kw) AND ((stroke* OR TIA OR acute NEXT coronar* NEXT syndrome* OR ((coronary) NEAR/3 (arter* OR heart*) NEAR/3 (disease*)) OR ((cardiovascular* OR cardiac*) NEAR/3 (event*)) OR bleeding* OR ((cerebrovascular) NEAR/3 (accident* OR disease*)) OR ischemi* OR (myocard* NEAR/3 infarct*)):ab,ti,kw) AND ((“CYP2C19” OR “CYP 2C19” OR “cytochrome P450 2C19”):ab,ti,kw)

**Google Scholar** *Top 200 relevant records*

clopidogrel|Plavix|P2Y12 stroke|ischemia|ischemic|TIA|"coronary artery|heart disease"|"cardiovascular|cardiac event"|bleeding|"cerebrovascular accident|disease"|"myocardial infarction" CYP2C19|"CYP 2C19"|"cytochrome P450 2C19"

**Supplementary Material II.** Cochrane Collaboration Risk of Bias assessment tool

### Domain 1: Risk of bias arising from the randomization process

| Signalling questions | Comments | Response options |
|---|---|---|
| **1.1 Was the allocation sequence random?** | | Y / PY / PN / N / NI |
| **1.2 Was the allocation sequence concealed until participants were enrolled and assigned to interventions?** | | Y / PY / PN / N / NI |
| **1.3 Did baseline differences between intervention groups suggest a problem with the randomization process?** | | Y / PY / PN / N / NI |
| **Risk-of-bias judgement** | | Low / High / Some concerns |
| Optional: What is the predicted direction of bias arising from the randomization process? | | NA / Favours experimental / Favours comparator / Towards null /Away from null / Unpredictable |

Domain 2: Risk of bias due to deviations from the intended interventions (*effect of assignment to intervention*)

| **Signalling questions** | **Comments** | **Response options** |
|---|---|---|
| **2.1. Were participants aware of their assigned intervention during the trial?** | | Y / PY / PN / N / NI |
| **2.2. Were carers and people delivering the interventions aware of participants' assigned intervention during the trial?** | | Y / PY / PN / N / NI |
| **2.3. If Y/PY/NI to 2.1 or 2.2: Were there deviations from the intended intervention that arose because of the trial context?** | | NA / Y / PY / PN / N / NI |
| **2.4 If Y/PY to 2.3: Were these deviations likely to have affected the outcome?** | | NA / Y / PY / PN / N / NI |
| **2.5. If Y/PY/NI to 2.4: Were these deviations from intended intervention balanced between groups?** | | NA / Y / PY / PN / N / NI |
| **2.6 Was an appropriate analysis used to estimate the effect of assignment to intervention?** | | Y / PY / PN / N / NI |
| **2.7 If N/PN/NI to 2.6: Was there potential for a substantial impact (on the result) of the failure to analyse participants in the group to which they were randomized?** | | NA / Y / PY / PN / N / NI |

| | | |
|---|---|---|
| **Risk-of-bias judgement** | | Low / High / Some concerns |
| Optional: What is the predicted direction of bias due to deviations from intended interventions? | | NA / Favours experimental / Favours comparator / Towards null /Away from null / Unpredictable |

Domain 3: Missing outcome data

| **Signalling questions** | **Comments** | **Response options** |
|---|---|---|
| **3.1 Were data for this outcome available for all, or nearly all, participants randomized?** | | Y / PY / PN / N / NI |
| **3.2 If N/PN/NI to 3.1: Is there evidence that the result was not biased by missing outcome data?** | | NA / Y / PY / PN / N |
| **3.3 If N/PN to 3.2: Could missingness in the outcome depend on its true value?** | | NA / Y / PY / PN / N / NI |
| **3.4 If Y/PY/NI to 3.3: Is it likely that missingness in the outcome depended on its true value?** | | NA / Y / PY / PN / N / NI |
| **Risk-of-bias judgement** | | Low / High / Some concerns |

| | | |
|---|---|---|
| Optional: What is the predicted direction of bias due to missing outcome data? | | NA / Favours experimental / Favours comparator / Towards null /Away from null / Unpredictable |

Domain 4: Risk of bias in measurement of the outcome

| **Signalling questions** | **Comments** | **Response options** |
|---|---|---|
| **4.1 Was the method of measuring the outcome inappropriate?** | | Y / PY / PN / N / NI |
| **4.2 Could measurement or ascertainment of the outcome have differed between intervention groups?** | | Y / PY / PN / N / NI |
| **4.3 If N/PN/NI to 4.1 and 4.2: Were outcome assessors aware of the intervention received by study participants?** | | NA / Y / PY / PN / N / NI |
| **4.4 If Y/PY/NI to 4.3: Could assessment of the outcome have been influenced by knowledge of intervention received?** | | NA / Y / PY / PN / N / NI |
| **4.5 If Y/PY/NI to 4.4: Is it likely that assessment of the outcome was influenced by knowledge of intervention received?** | | NA / Y / PY / PN / N / NI |

| | | |
|---|---|---|
| **Risk-of-bias judgement** | | Low / High / Some concerns |
| Optional: What is the predicted direction of bias in measurement of the outcome? | | NA / Favours experimental / Favours comparator / Towards null /Away from null / Unpredictable |

Domain 5: Risk of bias in selection of the reported result

| **Signalling questions** | **Comments** | **Response options** |
|---|---|---|
| **5.1 Were the data that produced this result analysed in accordance with a pre-specified analysis plan that was finalized before unblinded outcome data were available for analysis?** | | Y / PY / PN / N / NI |
| **Is the numerical result being assessed likely to have been selected, on the basis of the results, from...** | | |
| **5.3. ... multiple eligible outcome measurements (e.g. scales, definitions, time points) within the outcome domain?** | | Y / PY / PN / N / NI |
| **5.3. ... multiple eligible analyses of the data?** | | Y / PY / PN / N / NI |

| | | |
|---|---|---|
| **Risk-of-bias judgement** | | Low / High / Some concerns |
| Optional: What is the predicted direction of bias due to selection of the reported result? | | NA / Favours experimental / Favours comparator / Towards null /Away from null / Unpredictable |

Overall risk of bias

| | | |
|---|---|---|
| **Risk-of-bias judgement** | | Low / High / Some concerns |
| Optional: What is the overall predicted direction of bias for this outcome? | | NA / Favours experimental / Favours comparator / Towards null /Away from null / Unpredictable |

**Supplementary Table I.** PRISMA checklist

| Section/topic | # | Checklist item | Reported on page # |
|---|---|---|---|
| TITLE | | | |
| Title | 1 | Identify the report as a systematic review, meta-analysis, or both. | 1 |
| ABSTRACT | | | |
| Structured summary | 2 | Provide a structured summary including, as applicable: background; objectives; data sources; study eligibility criteria, participants, and interventions; study appraisal and synthesis methods; results; limitations; conclusions and implications of key findings; systematic review registration number. | 2, 3 |
| INTRODUCTION | | | |
| Rationale | 3 | Describe the rationale for the review in the context of what is already known. | 4, 5 |
| Objectives | 4 | Provide an explicit statement of questions being addressed with reference to participants, interventions, comparisons, outcomes, and study design (PICOS). | 5 |
| METHODS | | | |
| Protocol and registration | 5 | Indicate if a review protocol exists, if and where it can be accessed (e.g., Web address), and, if available, provide registration information including registration number. | ID: 242993 |
| Eligibility criteria | 6 | Specify study characteristics (e.g., PICOS, length of follow-up) and report characteristics (e.g., years considered, language, publication status) used as criteria for eligibility, giving rationale. | 5,6 |
| Information sources | 7 | Describe all information sources (e.g., databases with dates of coverage, contact with study authors to identify additional studies) in the search and date last searched. | 5 |
| Search | 8 | Present full electronic search strategy for at least one database, including any limits used, such that it could be repeated. | Supplement |
| Study selection | 9 | State the process for selecting studies (i.e., screening, eligibility, included in systematic review, and, if applicable, included in the meta-analysis). | 6 |
| Data collection process | 10 | Describe method of data extraction from reports (e.g., piloted forms, independently, in duplicate) and any processes for obtaining and confirming data from investigators. | 6 |
| Data items | 11 | List and define all variables for which data were sought (e.g., PICOS, funding sources) and any assumptions and simplifications made. | 5,6 |

| | | | |
|---|---|---|---|
| Risk of bias in individual studies | 12 | Describe methods used for assessing risk of bias of individual studies (including specification of whether this was done at the study or outcome level), and how this information is to be used in any data synthesis. | 6, 7 |
| Summary measures | 13 | State the principal summary measures (e.g., risk ratio, difference in means). | 7 |
| Synthesis of results | 14 | Describe the methods of handling data and combining results of studies, if done, including measures of consistency (e.g., $I^2$) for each meta-analysis. | 7 |

| Section/topic | # | Checklist item | Reported on page # |
|---|---|---|---|
| Risk of bias across studies | 15 | Specify any assessment of risk of bias that may affect the cumulative evidence (e.g., publication bias, selective reporting within studies). | 7 |
| Additional analyses | 16 | Describe methods of additional analyses (e.g., sensitivity or subgroup analyses, meta-regression), if done, indicating which were pre-specified. | 7 |
| **RESULTS** | | | |
| Study selection | 17 | Give numbers of studies screened, assessed for eligibility, and included in the review, with reasons for exclusions at each stage, ideally with a flow diagram. | 8 |
| Study characteristics | 18 | For each study, present characteristics for which data were extracted (e.g., study size, PICOS, follow-up period) and provide the citations. | 8 |
| Risk of bias within studies | 19 | Present data on risk of bias of each study and, if available, any outcome level assessment (see item 12). | 9 |
| Results of individual studies | 20 | For all outcomes considered (benefits or harms), present, for each study: (a) simple summary data for each intervention group (b) effect estimates and confidence intervals, ideally with a forest plot. | 9,10 |
| Synthesis of results | 21 | Present results of each meta-analysis done, including confidence intervals and measures of consistency. | 9,10 |
| Risk of bias across studies | 22 | Present results of any assessment of risk of bias across studies (see Item 15). | 9 |
| Additional analysis | 23 | Give results of additional analyses, if done (e.g., sensitivity or subgroup analyses, meta-regression [see Item 16]). | 9,10 |
| **DISCUSSION** | | | |
| Summary of evidence | 24 | Summarize the main findings including the strength of evidence for each main outcome; consider their relevance to key groups (e.g., healthcare providers, users, and policy makers). | 10,11 |
| Limitations | 25 | Discuss limitations at study and outcome level (e.g., risk of bias), and at review-level (e.g., incomplete retrieval of identified research, reporting bias). | 11,12 |

| | | | |
|---|---|---|---|
| Conclusions | 26 | Provide a general interpretation of the results in the context of other evidence, and implications for future research. | 12,13, 14 |
| FUNDING | | | |
| Funding | 27 | Describe sources of funding for the systematic review and other support (e.g., supply of data); role of funders for the systematic review. | Nvt |

*From:* Moher D, Liberati A, Tetzlaff J, Altman DG, The PRISMA Group (2009). Preferred Reporting Items for Systematic Reviews and Meta-Analyses: The PRISMA Statement. PLoS Med 6(7): e1000097. doi:10.1371/journal.pmed1000097

For more information, visit: **www.prisma-statement.org**.

**Supplementary Table II.** All poor and intermediate metabolizers of *CYP2C19* for each included study. *ASA = aspirin

| **Study** | **Intermediate metabolizers – Clopidogrel/ASA*** | **Poor metabolizers – Clopidogrel/ASA*** | **Intermediate metabolizers – ASA*** | **Poor metabolizers – ASA*** |
|---|---|---|---|---|
| **CHANCE** | 664 | 181 | 675 | 188 |
| **POINT** | 93 | 11 | 92 | 10 |
| **CURE** | 441 | 61 | 447 | 55 |
| **ACTIVE-A** | 95 | 10 | 94 | 12 |
| **CHARISMA** | 460 | 52 | 446 | 47 |
| **Yi et al.** | NI | NI | NI | NI |

**Supplementary Table III.** Investigated outcomes in each trial

| Study | Participants | Outcomes reported | Safety outcome | Major bleeding |
|---|---|---|---|---|
| CHANCE | Acute minor IS, TIA | Stroke, composite outcome (IS, hemorrhagic stroke, MI, vascular death) | Yes | Moderate or severe/life-threatening according to the GUSTO-criteria [18] |
| POINT | Acute minor IS, TIA | Composite outcome (IS, MI, ischemic vascular death) | Yes | Symptomatic intracranial hemorrhage, intraocular hemorrhage causing vision loss, trans- fusion of ≥2 units of red blood cells or an equivalent of whole blood, hospitalization or prolongation of an existing hospitalization, or death due to hemorrhage |
| CURE | ACS without ST-segment elevation | Composite outcome (IS, MI, ischemic vascular death) | Yes | Substantially disabling bleeding, intraocular bleeding leading to the loss of vision, or bleeding necessitating the transfusion of at least 2 units of blood |
| ACTIVE - A | AF | Major vascular event (Stroke, Embolism outside CNS, MI, vascular death) | Yes | Any overt bleeding requiring transfusion of at least two units of blood or meeting the criteria for severe hemorrhage. |
| CHARISMA | CAD, Multiple atherothrombotic risk factors, peripheral artery disease, cerebrovascular disease | Composite outcome (MI, Stroke, vascular death) | Yes | Moderate or severe/life-threatening according to the GUSTO-criteria[18] |
| Yi et al. | IS | Composite outcome (MI, IS, TIA, death) | No | - |

**Supplementary Table IV.** Cochrane Collaboration Risk of Bias assessment tool

| | CHANCE | POINT | Yi et al. | CURE | CHARISMA | ACTIVE-A |
|---|---|---|---|---|---|---|
| **Randomization process** | | | | | | |
| **Deviations from the intended interventions: effect of assignment of interventions** | | | | | | |
| **Deviations from the intended interventions: effect of adhering to interventions** | | | | | | |
| **Missing outcome data** | | | | | | |
| **Outcome measurement** | | | | | | |
| **Reported results** | | | | | | |
| **RoB Total Score** | **Low** | **Low** | **Some concern** | **Low** | **Low** | **Low** |
